# Clinical and neurobiologic predictors of long-term outcome in schizophrenia

**DOI:** 10.1101/2022.08.05.22278122

**Authors:** Thomas Nickl-Jockschat, Beng-Choon Ho, Nancy Andreasen

## Abstract

**Background:** Schizophrenia is a severe neuropsychiatric disorder accompanied by debilitating cognitive and psychosocial impairments over the course of the disease. As disease trajectories exhibit considerable inter-individual heterogeneity, early clinical and neurobiological predictors of long-term outcome are desirable for personalized treatment and care strategies.

**Methods:** In a naturalistic longitudinal approach, 381 schizophrenia patients from the Iowa Longitudinal Study (ILS) cohort underwent an extensive characterization, including repeated magnetic resonance imaging (MRI) scans, over a mean surveillance period of 11.07 years. We explored whether pre-diagnostic markers, clinical markers at the first psychotic episode, or magnetic resonance imaging (MRI) measures at the onset of the disease were predictive of relapse or remission of specific symptom patterns later in life.

**Results:** We identified a set of clinical parameters - namely premorbid adjustment during adolescence, symptom patterns, and neuropsychological profiles at disease onset – that were highly correlated with future disease trajectories. In general, brain measures *at baseline* did not correlate with outcome. *Progressive* regional brain volume losses over the observation period, however, were highly correlated with relapse patterns and symptom severity.

**Conclusions:** Our findings provide clinicians with a set of highly robust, easily acquirable, and cost-effective predictors for long-term outcome in schizophrenia. These results can be directly translated to a clinical setting to improve prospective care and treatment planning for schizophrenia patients. (Funding sources: NIH MH68380, MH31593, MH40856, and MH43271).

## Introduction

Schizophrenia is a debilitating neuropsychiatric disorder that typically begins during adolescence or young adulthood and leads to grave personal, familial, and socioeconomic consequences. Due to its early manifestation and comparatively high prevalence, schizophrenia is the 8th leading cause of disability-adjusted life years worldwide in the age group 15-44 years^1^. Symptoms are heterogeneous and are usually categorized in three groups: positive (or psychotic) symptoms, such as delusions and hallucinations, that occur most severely during psychotic episodes; negative symptoms, e.g., affective flattening, avolition and cognitive impairments, that often worsen during the course of the disease; and disorgnized symptoms, such as bizarre behavior^2^. Despite ongoing efforts to improve outcomes for these patients, many symptoms, especially negative symptoms, and cognitive impairments, remain largely nonresponsive to current pharmaceutical approaches.

When patients and their families are first confronted with this diagnosis, they frequently inquire about the prognosis. While there is consensus that affected individuals may face increasing cognitive and psychosocial deficits over the course of the disease^3^, clinicians and researchers also acknowledge a considerable heterogeneity of individual disease trajectories^4^. This heterogeneity confronts clinicians with several problems: first, prospective planning of care and treatment is significantly limited due to prognostic uncertainty; second, patients and their relatives naturally have high interest in how to plan the future; third, it is unknown whether this clinical diversity reflects different pathophysiologies, and if so, whether individual treatments could be optimized, when the approach is based on individual disease mechanisms.

The latter problem points to another unsolved riddle: whether clinical or neurobiological parameters are better suited for prognostic use. Neuroanatomical changes in schizophrenia are likely the best characterized biomarkers of this disease. Brain volume loss in a fronto-temporo-thalamo-basal ganglia network has been robustly identified in patients^5,6^. Importantly, these changes appear to be progressive throughout the disease course^5^ and are linked, at least in retrospective studies, to symptom severity^7^. However, there is a high heterogeneity of findings regarding regional brain volume decreases in patients^8^. While neuroanatomical heterogeneity renders neuroimaging measures unsuitable for diagnostic purposes, they might reflect the neurobiological basis for the heterogenous disease trajectories. However, it remains unknown whether neuro-anatomical changes can serve as predictors of disease outcomes. Thus, the quest for powerful predictors remains largely unsolved.

The Iowa Longitudinal Study (ILS) cohort was designed to create a unique data set to address these questions^9^. To our knowledge, it is the largest cohort that provides a long-term clinical and neurobiological characterization of patients, spanning an observation interval of nearly two decades, and providing frequent and regular clinical and biological measures. Hence, we relied upon this sample to identify clinical and neurobiological predictors of long-term outcome. In particular, we investigated three types of predictors which had been previously identified as potential biomarkers of schizophrenia: pre-diagnostic markers, clinical measures at the first psychotic episode, and neuroanatomical changes. Our findings provide clinicians with robust predictors of long-term outcome in schizophrenia, thus, filling a critical gap in the clinical care of these patients.

## Methods

### Patients and Assessments

We studied 381 schizophrenia patients (274 males and 107 females) drawn from the Iowa Longitudinal Study (ILS), based on the criterion that they underwent at least 2 years of surveillance (mean time of surveillance: 11.07 years, SD: 6.12 years). The surveillance period started with the first hospitalization or outpatient treatment for a schizophrenia spectrum disorder according to DSM-III and/or DSM/IV (see Supplement). At intake and every 6 months during the entire surveillance period, symptom severity and specificity were assessed and the treatment regimen was documented. Patients additionally received a structural MRI scan and cognitive testing at intake and at 2, 5, 9, 12, 15 and 18 years. A more detailed description of the ILS protocol can be found in Andreasen et al.^10^. Demographical and clinical characteristics of the sample are summarized in Table 1. For specific details on the acquisition parameters, please refer to the Supplement^5^.

**Table 1.** Overview over the demographic characteristics of the sample. Please note the distinction between the age at onset (i.e., the manifestation of any clinical symptoms that allow a diagnosis of schizophrenia) and the age at first psychotic episode (i.e., an episode with mainly positive symptoms). Duration of untreated psychosis (i.e., the time between the first psychotic episode and the initiation of antipsychotic treatment) was comparatively short. Times in remission/relapse are given as per cent of total time in surveillance. SD: standard deviation. A detailed summary of the distribution of these parameters across the three groups used in our GLM can be found in the Supplementary Table 1.

|  |  |
| --- | --- |
| **Sex ratio of the entire sample (males:females)** | 274:107 (71.92 per cent males, 28.08 per cent females) |
| **Years of education** | 12.57 (SD: 2.41) years |
| **Age at onset of disease** | 21.17 (SD: 6.31) years |
| **Age at first psychotic episode** | 23.58 (SD: 6.50) years |
| **Age at first medication** | 24.30 (SD: 7.09) years |
| **Age at first hospitalisation** | 24.41 (SD: 7.18) years |
| **Years between first psychotic episode and the initiation of pharmacological treatment (i.e., duration of untreated psychosis)** | 0.73 (SD: 2.63) years |
| **Age at the start of surveillance** | 24.16 (SD: 7.11) years |
| **Years in surveillance** | 11.07 (SD: 6.12) years |
| **Time in remission (per cent of total time in surveillance)** | 27.29 (SD: 31.97) |
| **Time in general relapse (per cent of total time in surveillance)** | 27.65 (SD: 27.90) |
| **Time in psychotic relapse (per cent of total time in surveillance)** | 13.71 (SD: 22.92) |
| **Time in negative relapse (per cent of total time in surveillance)** | 19.04 (SD: 24.55) |
| **Time in disorganized relapse (per cent of total time in surveillance)** | 2.54 (SD: 8.28) |

### MRI image analysis

The MRI data sets were analyzed with the AutoWorkup tool^11^, a fully automated image processing pipeline that is based on the BRAINS2 software package^12,13^. Further information on this workflow can be found in the Supplement.

### Statistical analyses: pre-diagnostic, clinical and neuropsychological measures

We used two different statistical approaches to evaluate potential predictors of long-term outcome. First, we grouped patients according to their disease trajectories in three groups: 1) patients that remitted after their first episode and never met relapse criteria (*remission only*), 2) patients that experienced both remission and relapse (*mixed*), and 3) patients that never experienced remission, but met relapse criteria (*relapse only*) over the entire observation period (please refer to Table 2 and the Supplement). We used a general linear model to evaluate whether pre-diagnostic markers, symptom patterns and neuropsychological markers *at onset* were associated with group status. This model allowed us to identify parameters at onset that were predictive of the general future disease trajectory.

**Table 2.** Prediagnostic parameters predict long-term relapse patterns. These parameters were assessed retrospectively with patients and their relatives during the first assessment upon inclusion into the study. Significant findings are highlighted in bold. Means are shown for the three subgroups of patients: “remission only” (patients that remitted after their first episode and never met relapse criteria), “relapse only” (patients that never experienced remission, but met relapse criteria), and “mixed” (patients that experienced both remission and relapse). Note that premorbid adjustment in childhood showed a strong trend, however, did not reach the significance threshold. Please also note that higher premorbid adjustment scores represent worse functioning.

|  | Mean Re-<br>mission | Mean<br>Relapse | Mean<br>Both | F | Pr>F | Duncan |
| --- | --- | --- | --- | --- | --- | --- |
| Age of onset | 21.58 | 20.83 | 21.35 | .39 | .6772 |  |
| Age at first psychotic medication | 23.88 | 23.02 | 23.99 | .98 | .3750 |  |
| Premorbid child Score | 2.61 | 3.50 | 3.30 | 3.01 | .0503 | 1(L) vs 2,3 |
| Premorbid adolescence Score | **3.74** | **5.68** | **5.04** | **9.22** | **.0001** | **1(L) vs 2,3** |
| Premorbid total Score | **6.30** | **9.19** | **8.31** | **7.15** | **.0009** | **1(L) vs 2,3** |
| Time between psychotic episode<br>and first medication | .62 | .81 | .67 | .16 | .8539 |  |

We, next, wanted to investigate, whether these parameters were not only suitable to predict relapse and remission *per se*, but were also possible to allow inference concerning future *symptom patterns*. We used parallel approaches with Pearson correlations to identify parameters at onset that were predictive of *distinct future symptom patterns*. We relied upon three-factor dimensions of psychotic, negative and disorganized symptoms^14^ to define remission or relapse during the observation period (Supplement). Patients were regarded to relapse in one of these dimensions (i.e., psychotic, negative or disorganized relapse) or in general, if respective symptoms were present for more than 2 weeks.

### Statistical analyses: regional brain volumes assessed by MRI

Schizophrenia is widely regarded to be a brain disease. When MR imaging was initially used to obtain brain measures in patients^15^, it was hoped that they would provide useful prognostic measures. Thus, the statistical analyses of our MRI data sets were motivated by two aims:

1. To determine whether variation in regional brain volumes *during the first episode* were correlated with time spent in relapse or remission *over the course of the disease*.
2. To determine whether changes in regional brain volumes *over the course of the disease* were correlated with time spent in remission or relapse.

For aim 1, we used the MRI data sets of our patients during first episode (i.e., the scan acquired at intake) to measure overall and regional brain volumes, which were normalized for intracranial volume. For aim 2, changes in regional brain volumes were defined as difference in volume between the first and the latest measurement in an individual subject, normalized by intracranial brain volume.

We then used General linear models to investigate whether regional brain volumes at onset (aim 1) or regional brain volume changes (aim 2) were associated with group status (remission only, relapse only and mixed), paralleling the statistical design of our analyses of clinical and neuropsychological parameters. Focusing on symptom patterns, we used a parallel design with Pearson correlations. To control for confounds, age at the time of the first MRI scan, gender, intracranial volume, MRI protocol, and dose years for antipsychotic medication^10^ were used as partial variables in each of the analysis approaches.

## Results

### Pre-diagnostic markers allow prediction of long-term outcome

Schizophrenia is increasingly recognized as a neurodevelopmental disorder with pathophysiological processes occurring long before the manifestation of the first psychotic episode^16-18^. There has been a strong interest in early diagnosis (i.e., identifying individuals who will later develop the full-blown illness), with the hope that early treatment might prevent its onset or reduce its severity^19^. Therefore, we aimed to explore whether premorbid features, such as poor social functioning, might indicate a more malignant pathophysiology and allow prediction of the future disease trajectory (Table 2).

Corroborating our initial hypothesis, pre-diagnostic markers were significantly associated with the later disease trajectory. Premorbid adjustment in adolescence was identified by our GLM analyses as the best predictor of future relapse status (p<.0001), followed by the premorbid total score (i.e., both childhood and adolescence premorbid scores combined) (p<.0009). Premorbid adjustment in childhood alone showed a strong trend, but failed to reach significance (p<.0503). Congruent with these findings, Pearson correlations for premorbid adjustment in adolescence were significant for time in general (r=.162; p=.002), negative (r=.169; p=.001) and disorganized (r=.117; p=.025) relapse. Adjustment scores in childhood were also associated with time in general relapse (p=.046), although the resulting p-values were higher. These results point to a manifestation of subtle deficits in adjustment as early as during childhood, getting more pronounced during adolescence (i.e., nearing disease onset).

To our surprise, none of the measures at or after the onset of symptoms was significantly correlated with long-term outcome. Specifically, time spent in first episode without treatment did not come close to significance in any of our analyses. Early intervention with conventional antipsychotic treatment strategies does not appear to positively influence the long-term in this sample.

### Symptom patterns and neuropsychological profiles at baseline correlate with long-term relapse and remission

We next evaluated whether the severity of distinct symptom clusters (Table 3) or the performance in standard neuropsychological tests during first episode (Table 4) were useful for predicting future relapse or remission. Strikingly, negative symptoms *at onset* stood out as the symptom cluster predicting future general relapse across our analyses. Of note, we found the severity of one symptom dimension at first assessment to be significantly correlated with the time spent in relapse for the same symptom dimension, but not for the other two dimensions. For example, psychotic symptoms at intake were significantly correlated with later time in psychotic relapse, but not in negative or disorganized relapse. Again, the lowest p-values for these associations was found for negative symptoms at baseline, which predicted the future burden of negative symptoms (p<.0001).

**Table 3.** Correlations of symptom clusters at baseline and time spent in relapse or remission later. Please note that the severity of one symptom cluster at first assessment was significantly associated with the time spent in relapse for the same symptom dimension, but not for the other two dimensions. Only negative symptoms were associated with time in general relapse. Significant correlations are highlighted in bold.

| Pearson Correlation Coefficients (Rho) with p-values |  |  |  |  |  |  |
| --- | --- | --- | --- | --- | --- | --- |
|  |  | Time in General Relapse | Time in Negative Relapse | Time in Psychotic Relapse | Time in Disorganized Relapse | Time in Remission |
| Positive symptoms score at baseline | Rho | **0.123** | 0.068 | **0.177** | 0.017 | -0.031 |
|  | *p* | **0.016** | 0.184 | **0.001** | 0.745 | 0.544 |
| Negative systems score at baseline | Rho | **0.256** | **0.311** | 0.095 | 0.068 | **-0.252** |
|  | *p* | **<.0001** | **<.0001** | 0.0632 | 0.1868 | **<.0001** |
| Disorganized systems score at baseline | Rho | 0.034 | 0.027 | 0.009 | **0.134** | -0.060 |
|  | *p* | 0.511 | 0.596 | 0.865 | **0.009** | 0.239 |

**Table 4.** Neuropsychological performance at baseline predicts later relapse patterns. Impairments in three major domains – IQ scores, verbal memory and attention – was significantly associated with long-term relapse. Performance IQ showed a trend, however, failed to reach significant. Significant finding are highlighted in bold.

|  | Mean Re-<br>mission | Mean<br>Relapse | Mean<br>Both | F | Pr>F | Duncan |
| --- | --- | --- | --- | --- | --- | --- |
| **Verbal IQ** | **95.94** | **89.85** | **93.13** | **4.63** | **.0104** | 2(L) vs 1(H) |
| **Performance IQ** | 93.54 | 88.21 | 91.73 | 3.01 | .0506 | 2(L) vs 1(H) |
| **Full Scale IQ** | **94.25** | **88.18** | **91.78** | **4.35** | **.0137** | 2(L) vs 1(H) |
| **Verbal Memory** | **-1.39** | **-1.72** | **-1.34** | **5.78** | **.0034** | 2 (L) vs 3(H), (M) |
| **Speed/Attention** | **-0.92** | **-1.62** | **-1.38** | **7.87** | **.0004** | 1(H) vs 2(L), (M) |
| **Problem Solving** | -0.94 | -1.47 | -1.27 | 3.86 | .0220 | 2(L) vs 1(H) |
| **Language** | -1.07 | -1.45 | -1.22 | 2.28 | .1038 |  |
| **Visuospatial Skills** | -0.85 | -1.13 | -1.13 | .80 | .4518 |  |
| **Motor Skills** | -0.93 | -0.71 | -0.87 | 1.09 | .3388 |  |

Neuropsychological testing during first episode revealed that impairments in three major domains – IQ scores, verbal memory and attention – predicted later time in general relapse. Regarding future symptom patterns, all these parameters were significantly correlated with negative relapse, while only speed and attention were significantly correlated with disorganized symptoms as another dimension of relapse. Thus, neuropsychological measures at intake can assist clinicians in counseling patients and families about the future course of the illness, even though they are not standard intake measures as of now.

### Relating long-term outcome to brain structure at first episode and brain volume loss over the course of the disease

Previous research has identified a fronto-temporal pattern of brain structural changes in schizophrenia patients^5^, but it has remained unclear whether brain structural changes *at first episode* can serve as predictors of long-term remission or relapse, and therefore assist clinicians in counseling patients and families. Remarkably this was the only approach in which our statistical models yielded contradictory results^20^. While our GLM analysis pointed to fronto-temporo-thalamic volume reductions as predictive for the future relapse status, Pearson correlations only reached significance for the volume of the right putamen and the right caudate with later time spent in disorganized relapse. We interpreted this as indicative for lacking robustness of these analyses. Opposed to these contradictory findings at baseline, both statistical approaches convergently yielded multiple significant relations retrospectively between volume loss in various cortical and subcortical fronto-temporal regions and long-term symptom patterns (Table 5). Nearly all findings were related to psychotic relapse, while progressive volume loss in only three regions was correlated with negative relapse: volume change of the frontal lobe, the temporal cerebrospinal fluid and ventricle-to-brain ratio (Supplementary Tables 2 and 3).

**Table 5.** Analysis of variance for regional brain volume loss over the observation interval and relapse patterns. Only significant findings are listed. For an overview over the results for all brain regions, as well as for our analyses of MRI measures at baseline, please refer to Supplementary Table 2 and 3.

|  | Mean Remis-<br>sion | Mean Re-<br>lapse | Mean Both | F | Pr>F |
| --- | --- | --- | --- | --- | --- |
| Cerebral White | .74 | -.16 | -.20 | 3.16 | .0445 |
| Cerebral CSF | .55 | 6.86 | 4.04 | 5.73 | .0038 |
| Cerebral Tissue | -.01 | -.35 | -.26 | 4.23 | .0158 |
| Frontal White | .62 | -.39 | -.32 | 3.70 | .0265 |
| Frontal CSF | 1.85 | 9.17 | 5.19 | 5.69 | .0040 |
| Frontal Tissue | .00 | -.55 | -.40 | 6.13 | .0026 |
| Temporal CSF | -.21 | 5.76 | 3.93 | 5.13 | .0067 |
| Parietal Tissue | .09 | -.32 | -.27 | 3.88 | .0223 |
| Ventricle/Brain Ratio | -1.32 | 4.24 | 1.86 | 8.92 | .0002 |
| Left Putamen | -.91 | .07 | .01 | 3.74 | .0254 |

## Discussion

Schizophrenia is usually a serious life-long disorder. Hence, identifying factors at early stages of the disease that may predict long-term outcome is critical to improving the planning of treatment and care for patients. Our study identified three major predictors of disease trajectories: premorbid adjustment during adolescence, symptom patterns, and neuropsychological profiles at first hospitalization. These results provide clinicians with a set of parameters that are easy and cost-effective to acquire, and that allow robust inference of long-term prognosis. They also highlight the importance of subtle disease processes in affected individuals long before the actual manifestation of symptoms, which are only partially addressed by current treatment strategies.

Our finding that symptom clusters at onset were associated with later relapse patterns offers a unique opportunity for prognostic purposes. Particularly negative symptoms at the onset of the disorder were associated with future relapse and, hence, a more malignant course of the disorder. This complements previous studies that have highlighted the detrimental nature of negative symptoms, as these are strongly associated with deficits in various independent living skills and global functional deficits^20-22^, rendering negative symptoms as a key player in schizophrenia-associated disability^3,20,23^. Unfortunately, the pivotal impact of negative symptoms on global outcome measures is further aggravated by their minimal response to pharmacological interventions^24^. These results support a stronger research focus specifically on negative symptoms^24,25^. Clinicians should thoroughly assess this symptom dimension in their first-episode patients; at present, it tends to be neglected even by experienced providers^22^.

While only negative symptoms were predictive of future general relapse or remission, our finding that the severity of a certain symptom cluster at first assessment was significantly associated with the time spent in relapse later for the same symptom dimension offers a unique opportunity for an even more sophisticated prognosis. Clinical symptoms at first episode alone can serve as a valid predictor of later recurrence of the same symptoms. This opposes the common notion that symptom patterns in schizophrenia are too unstable to allow robust and clinically useful predictions^26,27^.

Neuropsychological parameters at first episode were also robust predictors of future time in remission and the burden of negative symptoms. Impairments in these domains are typically perceived as hallmarks of schizophrenia, namely attention^28,29^ and IQ^30,31^. Our finding suggests that first-episode patients may benefit from baseline neuropsychological testing, although this is not currently recommended as standard of care.

These results also offer important insight into the underlying neurobiology of the disorder. Schizophrenia is increasingly perceived as a neurodevelopmental disorder, with pathophysiological processes and subtle cognitive anomalies progressing long before the manifestation of clinical symptoms^16-18^. Neuroplastic changes during adolescence or early adulthood might lead to the emergence of first psychotic symptoms^16,32^. Congruent with these hypotheses, we found premorbid adjustment in adolescence as the best pre-diagnostic predictor of relapse, while adjustment scores during childhood still trended towards significance. We interpreted these results as indicators of early disturbed trajectories and a rapidly accelerated decline during adolescence. Such an interpretation is corroborated by cognitive impairments in children and adolescents that later present with schizophrenia^33,34^.

Of note, clinical and neuropsychological parameters were much better predictors of remission or relapse than much more costly and laborious analyses of neuroanatomical parameters at first episode. The advent of MR imaging has nurtured hopes that direct observations of brain pathologies could provide much better prediction of disease trajectories than clinical measures^35,36^. However, in this comprehensive longitudinal study, neuroanatomical markers at first episode did not appear to be useful for predicting long-term relapse or remission, as our statistical approaches failed to yield convergent results. This lack of convergence might be driven by two factors: first, although brain structural changes can be detected already in first episode patients^5^ and even prodromal individuals^37^, they still might be too subtle to be already suitable for prognostic purposes in MRI studies at baseline. Second, *progressive volume loss* rather than *absolute changes* in regional brain volumes of patients might be relevant for the emergence of symptoms and distinct disease trajectories. As baseline MRI is certainly unable to capture dynamic changes in brain structure, it might be less useful for long-term prediction.Our findings of a retrospective association between longitudinal brain volume changes in fronto-temporo-thalamic regions and psychopathology confirms earlier similar finding in this cohort^5,7^, but also in independent samples^6,38^.

While our findings render doubt on the value of an MRI scan at first episode for predictive purposes, we would like to emphasize that its use for differential diagnostic purposes, i.e., the detection of etiologically relevant (e.g., malignant, inflamatory or ischemic) brain lesions, remains unchallenged. Given their close relationship to distinct symptom clusters, progressive brain volume changes, in turn, appear to offer a major avenue towards a better understanding of the neurobiology of schizophrenia.

## Limitations

Although we employed a sophisticated statistical design to tease apart complex causal relationships, these cannot be completely disentangled. Such considerations are especially important for highly complex interactions between clinical symptoms and neuroanatomy. Cumulative doses of antipsychotic medication, for example, have been factored into our model as covariates, however, such a correction will not be able to account for all effects of this heterogeneous substance class.

The naturalistic design of this study entails other limitations. While a randomized controlled trial would have standardized treatment, it would have been impossible to implement such an approach over a prolonged observation period. On the other hand, a naturalistic approach pictures long-term trajectories much closer to clinical reality than a standardized trial.

## Conclusion

In summary, our findings provide a set of highly robust and easily acquirable predictors for longterm outcome in schizophrenia. Future neurobiological and treatment studies can capitalize on these predictors to further stratify patient populations and reduce heterogeneity accordingly.

## Data Availability

All data produced in the present study are stored at the University of Iowa.

