## Supplement for "Clinical and neurobiologic predictors of long-term outcome in schizophrenia"

### Methods

*Patients and Assessments*

381 schizophrenia patients (274 males and 107 females) drawn from the Iowa Longitudinal Study (ILS) were included in this study, if they underwent at least 2 years of surveillance. Diagnosis at intake was based on the Comprehensive Assessment of Symptoms and History (CASH), a structured interview developed for longitudinal research (Andreasen et al., 1992). The CASH interview is based on the diagnostic criteria of the Diagnostic and Stastical Manual of Mental Disorders. As recruitment for this cohort started in 1987 and ended in 2007, diagnoses referred to the actual DSM versions at the time of recruitment, i.e., DSM-III and IV. Exclusion criteria for this study included an IQ<70, a history of head trauma and general contraindications for MRI scans. Patients underwent clinical assessments at intake and every 6 months, during which symptom severity and specifity, psychosocial functions and the treatment regimen were documented across the entire timeline of surveillance. Also at intake and and 2, 5, 9, 12, 15 and 18 years after inclusion, patients received a structural MRI (sMRI) scan and an extensive cognitive testing.

*Clinical measures and definitions*

We used the well-established three-factor dimensions of positive, negative and disorganized symptoms (Andreasen et al., 1995) to define remission or relapse during the observation period. In detail, we relied upon the following definitions:

*Remission:* a period of time lasting at least 6 months, when none of the following symptoms were rated above 2 (mild) on the SAPS/SANS: delusions, hallucinations, bizarre behavior, positive formal thought disorder, alogia, avolition, and affective flattening. Note that the symptoms of interest do not include the following: anhedonia, attention or inappropriate affect. Brief period of mild symptoms were allowed, if they did not occur longer than 2 weeks and the remission resumed at least for a total of 6 months.

*Psychotic Relapse*: A rating of 4-5 (severe or extreme) on the SAPS/SANS for either hallucinations or delusions lasting continuously for at least 2 weeks.

*Negative Relapse*: A rating of 4-5 (severe or extreme) for at least two of the following: alogia, anhedonia, affective flattening, avolition, or attention lasting continuously for at least 2 weeks.

*Disorganized Relapse*: A rating of 4-5 (severe or extreme) on at least 1 of the following: positive formal thought disorder (PFTD), bizarre behavior, or inappropriate affect lasting continuously for at least 2 weeks.

*General Relapse*: Any period of time when any of the above types of relapse occured. A relapse ended when the symptoms were not rated as severe or extreme (4-5) for at least 4 weeks.

*Structural MRI data acquisition*

We used two scanning protocols that we refer to as MR5 and MR6 (Andreasen et al., 2011). Both are multimodal, as they acquire T2 or proton density (PD) sequences apart from T1, to provide optimal discrimination between grey and white matter. Each participant's data included T1-, T2-

, and PD-weighted images collected on a 1.5-T GE Signa scanner for MR5 scanning. MR6 scanning was performed on a 1.5-T Siemens Avanto scanner using T1and T2 sequences. Subjects continued in the same sequence throughout the study. In other words, those who began with an MR5 protocol remained in it for all scans, and those who began with an MR6 protocol remained in it as well. The two sequences differ primarily in slice thickness and in-plane resolution; both are acquired in the coronal plane. Voxel sizes for MR5 are 1.0×1.0×1.5 mm for T1 scans and 1.0×1.0×3 mm for T2 scans; voxel sizes for MR6 are 0.625×0.625×1.5 mm for T1 scans and 0.625×0.625×1.8 mm for T2 scans.

*MRI image analysis*

The MRI data sets were analyzed with the AutoWorkup tool (Pierson et al., 2011), a fully automated image processing pipeline that is based on the BRAINS2 software package (Andreasen et al., 1992; Magnotta et al., 2003). In brief, the analysis is composed of four major steps:

1) Realignment and coregistration of all scans into the same orientation and resolution
2) Tissue classification
3) Definition of the borders of the brain and parcellation into subregions and structures
4) Measurement of the volumes of brain regions and various tissues within predefined borders

This approach has been optimized for the assessment of brain structural changes in longitudinal samples (Pierson et al., 2011; Andreasen et al., 2013).

**Supplementary Table 1**

| | Remission Only | Mixed | Relapse Only |
|---|---|---|---|
| **Sex (M/F)** | 28/16 | 118/61 | 128/30 |
| **Age of surveillance (years)** | 24.01 (SD: 9.125) | 24.53 (SD: 6.943) | 23.78 (SD: 6.666) |
| **Surveillance Duration Total (years)** | 7.405 (SD: 4.476) | 12.31 (SD: 5.899) | 10.68 (SD: 6.322) |
| **Age of Onset (years)** | 21.59 (SD: 8.157) | 21.36 (SD: 6.439) | 20.84 (SD: 5.556) |
| **Age first psychotic episode (years)** | 23.89 (SD: 8.626) | 24.00 (SD: 6.628) | 23.02 (SD: 5.658) |
| **Age at first medication (years)** | 24.65 (SD: 9.204) | 24.55 (SD: 6.924) | 23.94 (SD: 6.686) |
| **Age at first hospitalization (years)** | 24.57 (SD: 9.584) | 24.60 (SD: 6.858) | 24.16 (SD: 6.899) |
| **Time between 1st psychotic symptoms and 1st medication (years)** | 0.626 (SD: 1.523) | 0.671 (SD: 2.854) | 0.817 (SD: 2.594) |

**Supplementary Table 1.** Distribution of demographic characteristics over the three groups (remission only, relapse only and mixed) used in our GLM analyses. The exact criteria that were used to assign patients to one of the groups are depicted in the methods part of the manuscript and in the supplement (above). Please note the distinction between the age at onset (i.e., the manifestation of any clinical symptoms that allow a diagnosis of schizophrenia) and the age at first psychotic episode (i.e., an episode with mainly positive symptoms). SD: standard deviation.

**Supplementary Table 2**

| | Mean Remission | Mean Relapse | Mean Both | p-value |
|---|---|---|---|---|
| Cerebral Gray | 690.37 | 700.51 | 682.95 | .1260 |
| Cerebral White | 445.21 | 462.25 | 444.28 | .1664 |
| **Cerebral CSF** | **66.12** | **73.77** | **84.14** | **.0070** |
| **Cerebral Tissue** | **1135.57** | **1162.76** | **1127.23** | **.0072** |
| Frontal Gray | 268.54 | 271.13 | 264.35 | .1185 |
| Frontal White | 180.78 | 187.07 | 180.67 | .3955 |
| **Frontal CSF** | **32.36** | **36.55** | **43.16** | **.0119** |
| Frontal Tissue | 449.32 | 458.20 | 445.02 | .0929 |
| Temporal Gray | 161.40 | 162.66 | 159.11 | .0730 |
| Temporal White | 70.78 | 73.49 | 70.32 | .1797 |
| **Temporal CSF** | **9.32** | **10.01** | **10.78** | **.0283** |
| Temporal Tissue | 232.18 | 236.16 | 229.43 | .0607 |
| Parietal Gray | 139.49 | 141.83 | 138.46 | .6266 |
| Parietal White | 104.13 | 109.80 | 105.06 | .0936 |
| Parietal CSF | 12.44 | 14.12 | 15.92 | .0718 |
| Parietal Tissue | 243.62 | 251.64 | 243.52 | .1727 |
| Occipital Gray | 69.69 | 72.16 | 69.46 | .2621 |
| Occipital White | 43.16 | 45.19 | 43.84 | .9442 |
| Occipital CSF | 2.19 | 2.37 | 2.45 | .7316 |
| Occipital Tissue | 112.85 | 117.35 | 113.30 | .3120 |
| Cortical Gray | 640.00 | 648.85 | 632.47 | .1190 |
| Cortical White | 399.20 | 415.95 | 400.35 | .2319 |
| **Cortical CSF** | **40.98** | **45.48** | **52.66** | **.0317** |
| Cerebellum | 132.68 | 135.98 | 133.14 | .8918 |
| **Ventricle/Brain Ratio** | **.99** | **1.18** | **1.39** | **.0032** |
| Right Putamen | 5.50 | 5.62 | 5.43 | .3813 |
| Left Putamen | 5.16 | 5.32 | 5.15 | .7463 |
| **Right Thalamus** | **6.33** | **6.25** | **6.04** | **.0023** |
| **Left Thalamus** | **6.75** | **6.71** | **6.42** | **.0007** |
| Right Caudate | 3.81 | 3.99 | 3.87 | .4949 |
| Left Caudate | 3.77 | 3.99 | 3.83 | .3021 |
| Right Hippocampus | 2.36 | 2.28 | 2.23 | .0997 |
| Left Hippocampus | 2.27 | 2.24 | 2.21 | .4357 |
| **Thalamus** | **13.08** | **12.96** | **12.46** | **.0008** |
| Putamen | 10.65 | 10.93 | 10.58 | .5547 |
| Caudate | 4.64 | 4.77 | 4.65 | .9751 |
| Hippocampus | 4.63 | 4.52 | 4.44 | .1820 |

**Supplementary Table 2.** MRI measures across the three groups (remission only, relapse only and mixed) at intake. Shown here are the volumes of all brain regions in our analyses and the p-values for potential differences between the three groups. Significant findings are displayed in bold.

**Supplementary Table 3**

| | Mean Remission | Mean Relapse | Mean Both | p-value |
|---|---|---|---|---|
| Cerebral Gray | -.48 | -.45 | -.27 | .2186 |
| **Cerebral White** | **.74** | **-.16** | **-.20** | **.0445** |
| **Cerebral CSF** | **.55** | **6.86** | **4.04** | **.0038** |
| **Cerebral Tissue** | **-.01** | **-.35** | **-.26** | **.0158** |
| Frontal Gray | -.39 | -.62 | -.43 | .1197 |
| **Frontal White** | **.62** | **-.39** | **-.32** | **.0265** |
| **Frontal CSF** | **1.85** | **9.17** | **5.19** | **.0040** |
| **Frontal Tissue** | **.00** | **-.55** | **-.40** | **.0026** |
| Temporal Gray | -.33 | -.28 | -.19 | .8089 |
| Temporal White | .46 | .01 | -.04 | .8251 |
| **Temporal CSF** | **-.21** | **5.76** | **3.93** | **.0067** |
| Temporal Tissue | -.10 | -.21 | -.16 | .6897 |
| Parietal Gray | -.25 | -.57 | -.37 | .0635 |
| Parietal White | .60 | .03 | -.10 | .2508 |
| Parietal CSF | .83 | 7.54 | 4.79 | .1843 |
| **Parietal Tissue** | **.09** | **-.32** | **-.27** | **.0223** |
| Occipital Gray | -.39 | -.09 | .34 | .0956 |
| Occipital White | 1.05 | .23 | -.36 | .0790 |
| Occipital CSF | .62 | 19.27 | 10.72 | .5763 |
| Occipital Tissue | -.00 | -.02 | -.02 | .9498 |
| Cortical Gray | -.37 | -.47 | -.28 | .0924 |
| Cortical White | .61 | -.15 | -.23 | .1025 |
| Cortical CSF | 2.03 | 8.32 | 5.82 | .0570 |
| Cerebellum | .18 | .02 | -.02 | .4796 |
| **Ventricle/Brain Ratio** | **-1.32** | **4.24** | **1.86** | **.0002** |
| Right Putamen | -.19 | .13 | .02 | .2969 |
| **Left Putamen** | **-.91** | **.07** | **.01** | **.0254** |
| Right Thalamus | -.82 | -.45 | -.26 | .1310 |
| Left Thalamus | -.82 | -.83 | -.55 | .4111 |
| Right Caudate | -.30 | .11 | .03 | .2104 |
| Left Caudate | .05 | -.04 | .12 | .5032 |
| Right Hippocampus | -.10 | .25 | .20 | .5629 |
| Left Hippocampus | .28 | .17 | -.02 | .8444 |
| Thalamus | -.98 | -.60 | -.39 | .1098 |
| Putamen | -.58 | .15 | .06 | .0609 |
| Caudate | -.61 | -.37 | -.13 | .1900 |
| Hippocampus | .08 | .18 | .08 | .9273 |

**Supplementary Table 3.** Brain volume loss across the three groups (remission only, relapse only and mixed) at intake. Shown here is the change of volume between the first measurement at intake and the last MRI of the study of all brain regions in our analyses and the p-values for potential differences between the three groups. Significant findings are displayed in bold.
